# Re-emergence of COVID-19 in Beijing was triggered by frozen virus: evidence from molecular clock

**DOI:** 10.1101/2021.04.19.21255726

**Authors:** Yunjun Zhang, Yong Zhang, Mifang Liang, Yi Zhang, Yang Li, Xuejun Ma, Xiaohua Zhou

## Abstract

The COVID-19 outbreak in Xinfadi (XFD) Wholesale market in Beijing, China in June, 2020 caused 368 reported cases within 39 days. Genetic evidences indicated that imported SARS-CoV-2 (belong to the lineage B1.1.29) initiated this outbreak. However, the transmission route of the virus is still unknown. We obtained from public database three SARS-CoV-2 genomes isolated in XFD (XFD genomes) and adopted the leaf-dating method to calculate their expected collection dates using temporal calibrating information from other 241 genomes collected in mainland of China. All three XFD genomes were calculated to have earlier collection dates than the recorded (Bayes factor >1), and hence exhibited a lack of genetic divergence. We additionally combined the XFD genomes with other 225 genomes subsampled from those of the lineage B1.1.29, among which five sequences were also included for control analysis. Two of three XFD genomes were calculated to have earlier collection dates than the recorded (Bayes factor >1), while no control genomes provided such evidence. According to present understanding of SARS-CoV-2, a lack of genetic divergence is most likely due to being frozen. Considering the fact that the XFD outbreak started from a booth of frozen food, we judged that the XFD outbreak was caused by contaminated frozen food. Our results provided molecular evidence for the source of COVID-19 outbreak in Beijing XFD, which highlights new targets for SARS-CoV-2 surveillance for the public health.

## INTRODUCTION

A COVID-19 case was identified in Beijing on 2020-06-11, ending a run of 55 days without local transmission and starting the second outbreak. Since then, the city has responded quickly and fiercely with a series of control measures, such as large scale qRT-PCR screening, locking down affected residential compounds, and closing all schools [1]. With all these efforts, the second outbreak was taken under control within two weeks, and lasted only for 39 days (till 2020-07-20) with 368 qRT-PCR positive cases [2].

Most cases of the outbreak had a history of close contact with Xinfadi (XFD) Wholesale Market. Genetic tracing of SARS-CoV-2 genomes from XFD indicated that the outbreak was triggered by imported virus. Specifically it was a member of the lineage B1.1 (now of the lineage B1.1.29, [3]) predominately circulating in Europe rather than in China [4]. However, how the virus was transmitted to Beijing has not been clarified. Evidences from epidemiological field investigation showed that a salmon booth (#S14) was the starting point [2], suggesting that the viruses could have been imported with contaminated frozen food. However, some held a belief that the virus was imported by infected human [1].

Molecular clock-based evolutionary analysis of temporally spaced genome data provides a new tool for timing past events and exploring the mechanism and process of evolution [5]. Generally, the molecular clock posits that viral genomes accumulate mutations at roughly a constant rate, and the process could be affected by certain environmental factors [6]. For example, frozen viral isolates do not accumulate genetic mutations while in storage [7]. This environmental factor, frozen in storage, could help test the contaminated seafood hypothesis. We therefore performed evolutionary analysis of the SARS-CoV-2 genome data of the isolates collected in the XFD Wholesale Market (denoted as XFD genomes). We evaluated the amount of genetic changes embedded in these genomes to test whether the contaminated seafood hypothesis worded for the import of virus leading to the second outbreak of COVID-19 in Beijing in 2020.

## RESULTS

Three human-source SARS-CoV-2 genomes from the XFD outbreak were obtained from open database of GISAID ([8], accessed on 2021-01-30). Assuming that XFD genomes follow a certain evolutionary rate, the expected collection dates (denoted as DateE) of XFD genomes were calculated with leaf-dating method based on the molecular clock.The virus founded in XFD outbreak belonged to the B1.1.29 lineage which was previously circulating outside of China. The rates were therefore generated with two datasets, one consisted of SARS-CoV-2 genomes sampled from the mainland of China (referred to as domestic dataset) and the other of the B1.1.29 lineage regardless of the region (referred to as lineage-specified dataset). Control analysis was conducted with five randomly selected SARS-CoV-2 genomes (referred to as control genomes) from the lineage of B1.1.29.

### XFD genomes

Based on the domestic dataset, all three XFD genomes were expected to have earlier collection dates than their recorded collection (Fig 1). We calculated that the XFD genome IVDC-02-06 was sampled on 2020-03-11 (95%HPD: 2020-01-21, 2020-05-08), being 92 days earlier than its recorded collection on 2020-06-11. The consequent Bayes foctor was 11.27, suggesting strong evidence in favour of the hypothesis (H_2_: Date_E_ ≠Date_r_, defined in the Fig 1) that the expected collection date of the genome is unequal to the recorded date.

**Figure 1.**
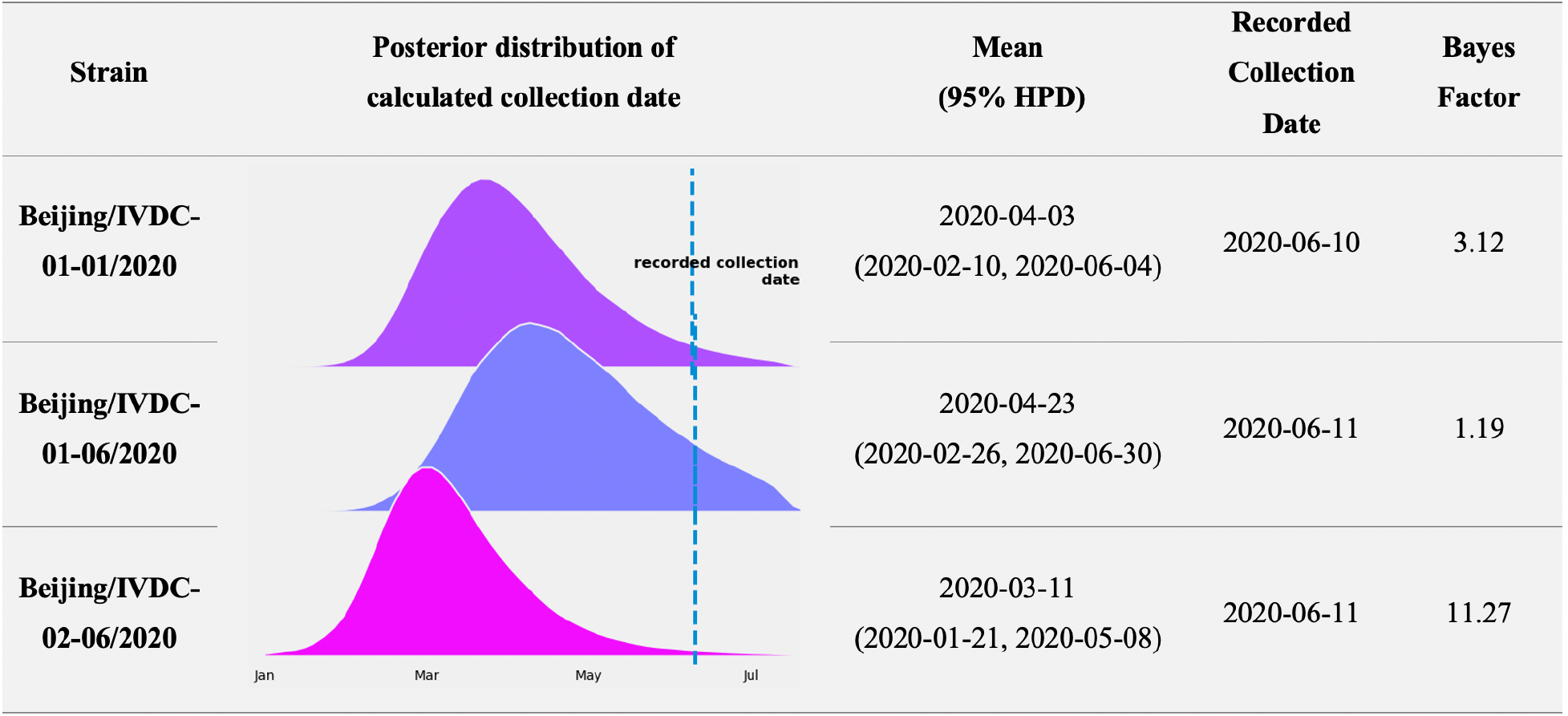
Calculated and recorded collection dates of XFD genomes based on the domestic dataset. Bayes factor is defined as the marginal likelihood of 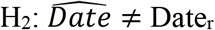 over that of 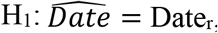, where 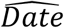 is the calculated collection date, and Date_r_ is the recorded collection date of a particular genome. The bayes factor exceeds 1, 3, or 10, suggesting weak, moderate, and strong evidence in favor of H_2_, respectively.

The other two genomes, IVDC-01-01 and IVDC-01-06 were calculated to be 68 and 49 days earlier than their recorded collections, respectively, providing moderate and weak level of bayes factor in favour of the hypothesis H_2_. Meanwhile, the evolutionary rate of SARS-CoV-2 genome was estimated to be 5.94× 10−4 substitutions per site per year (95% HPD: 3.94 × 10−4, 7.85 × 10−4), which was consistent with the previous study [9].

**Figure 1 Calculated and recorded collection dates of XFD genomes based on the domestic dataset.** Bayes factor is defined as the marginal likelihood of H_2_: Date_E_≠ Date_r_ over that of H_1_: Date_E_ = Date_r_, where Date_E_ is the expected collection date, and Dater the recorded collection date of a particular genome. The bayes factor exceeds 1, 3, or 10, suggesting weak, moderate, and strong evidence in favor of H_2_, respectively.

Based on the lineage-specified dataset, two XFD genomes (IVDC-01-01 and IVDC-02-06) provided weak evidence in favor of hypothesis H_2_ with the bayes factor 2.98 and 1.07, respectively (Fig 2). Compared with the results obtained from the domestic dataset, the calculated collection dates of the two XFD genomes were 11∼17 days later (but still earlier than the recorded collection dates) and with broader 95%HPD.

**Figure 2.**
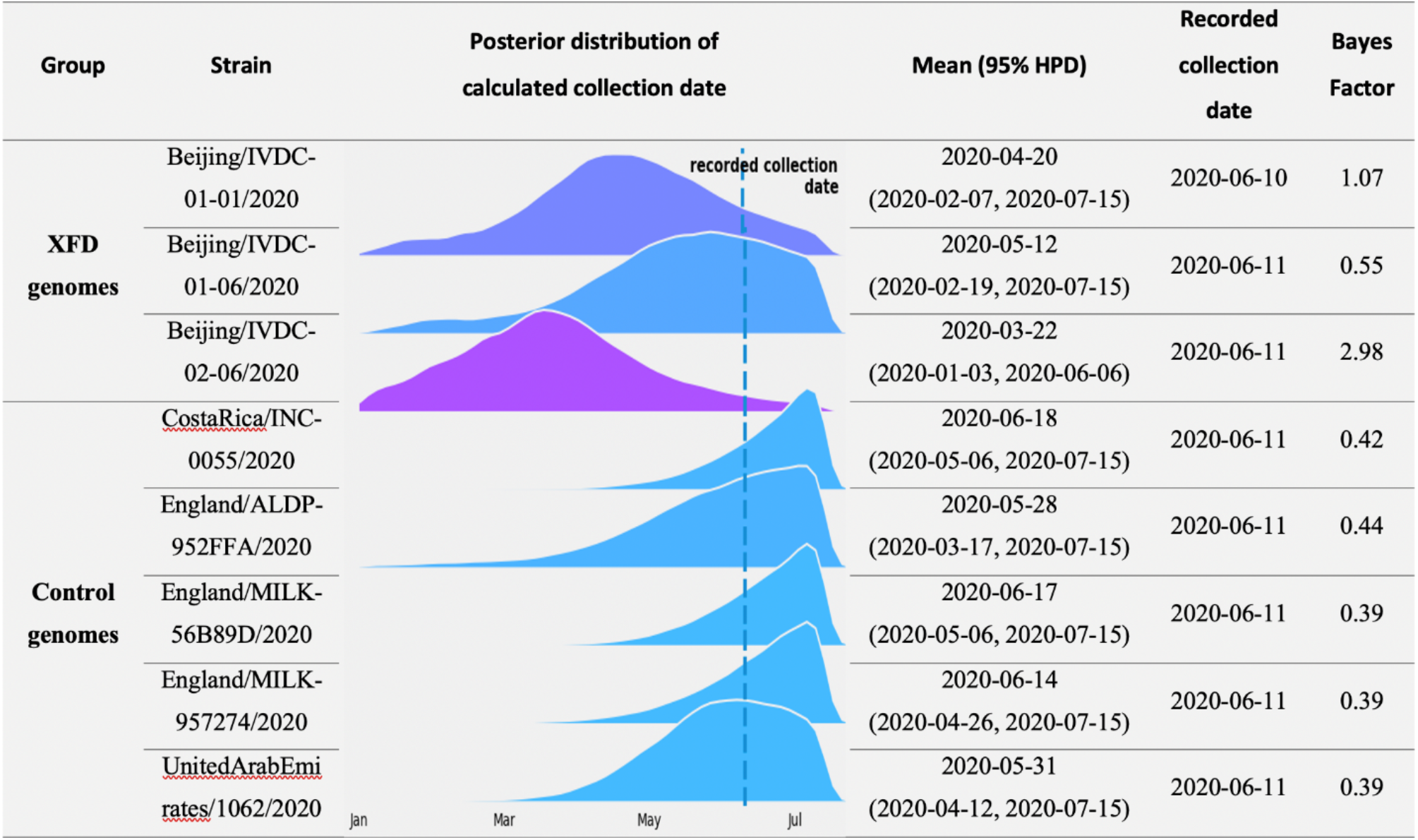
**Calculated and recorded collection dates of XFD genomes and control genomes based on the lineage-specified dataset**. The bayes factor suggests weak, moderate, and strong evidence in favor of H_1_ if being smaller than 1, 1/3, or 1/10, respectively.

Meanwhile, the evolutionary rate of SARS-CoV-2 genome was estimated to be 3.17× 10−4 substitutions per site per year (95% HPD: 2.38 × 10−4, 4.02 × 10−4), which was overlapped with the estimation from domestic dataset.

### Control genomes

In addition, the expected collection dates of the five control genomes were simultaneously calculated on the basis of lineage-specific dataset (Fig 2). Compared with those of the XFD genomes, the calculated collection dates of control genomes were consistently later and closer to their recorded collection dates, leading to consistently smaller bayes factors and weak evidence in favor of the hypothesis (H_1_:Date_E_ = Date_r_) that the expected collection date of the genome is equal to its recorded date.

**Figure 2 Calculated and recorded collection dates of XFD genomes and control genomes based on the lineage-specified dataset**. The bayes factor suggests weak, moderate, and strong evidence in favor of H_1_ if it is smaller than 1, 1/3, or 1/10, respectively.

## DISCUSSIONS

The evolutionary analysis of SARS-CoV-2 genomes sampled in XFD revealed that the two XFD genomes (IVDC-02-06 and IVDC-01-1) lacked genetic divergence. If they followed the previous evolutionary rate, the calculated collection date of these two XFD genomes were earlier than the record date.

This lack of genetic divergence was attributed to frozen storage for the following reasons. First, SARS-CoV-2 biology suggests it be impossible for some viruses staying in a host or reservoir for several months with little or no replication before reactivation and a second outbreak [6].

Second, it has been reported that SARS-CoV-2 can survive the time and temperatures associated with transportation and storage conditions associated with international food trade [10]. Besides, epidemiological evidence revealed that the outbreak started from a booth of frozen food in the market [2]. Therefore, we hypothesized that the contaminated imported food carried the live virus from the supply chain, survived in the frozen storage, and transferred the virus to the XFD market, thus leading the outbreak.

In addition, when analyzing the control genomes from lineage B1.1.29 with the same collection date of 2020-06-11 as that of the two XFD genomes, their calculated collection dates skewed to the right (Fig 2). Four of five genomes had the peak (of the posterior distribution of the calculated collection date) later than the acture collection date. This bias arise from the used leaf-dating method which exhibited moderate bias when the true age was small [7]. But when this method was applied to the XFD genomes, they showed peaks clearly ahead of their recorded collection dates. This further supported the XFD genomes being earlier position in the evolutionary process of SARS-CoV-2.

A real-world support was from a genome collected in Singapore (Singapore/487/2020 (EPI_ISL_476821)), which was identical to the XFD genome IVDC-02-06. The Singapore genome was collected from an international traveler on 2020-03-26, which was very close to our calculation (2020-03-22) of the XFD IVDC-02-06 with leaf-dating method. This lends proof to the validity of our estimation of collection date and provids further evidence that the genome should have appeared much earlier than the recorded 2020-06-11.

Previous studies have attributed the recurrence of infectious diseases to frozen viruses via phylogenetic and evolutionary analysis. Previous researches were based on either the phylogenetic clustering of viral genomes [11] or on the discontinuity of the evolutionary rates between the first and second outbreaks [6]. In this study, the leaf-dating method was used to estimate the collection time, and the Bayes factor was used to explore the gap between the actual collection time and the calculated collection time. An quantitative measure of the evidence for whether the virus being frozen and, if so, a composite estimation of the time length of being frozen were provided by our method. In addition, our method required less genome data from the second outbreak and could be performed independently for each genome collected in the second outbreak.

The conculsion of our study was made based on a relative homogeneous model setting: i.e., assuming the evolutionary rate be constant (strict molecular clock) and using the coalescent-based tree prior regardless of population structure. In recent years, it is more appealing to incorporate heterogeneity both in the molecular clock, such as relaxed molecular clock, and in the tree prior model, such as structured coalescent model. However, a previous study found that very similar estimation of collection date would be obtained under the strict or relaxed molecular model [7]. Hence our evidence for XFD genomes may not have been affected if adopting the relaxed molecular clock model. In addition, incorporating population structure into the tree prior model was demonstrated to considerably decrease the estimation of evolutionary rate [12]. This effect of a slower rate, had we adopted population structure, would have strengthened our evidence for earlier expected collection date.

In conclusion, we conclude that the outbreak in XFD market in Beijing was caused by contaminated frozen food (or its packaging). Our findings, coupled with the reports from China of SARS-CoV-2 being detected on imported frozen food, should alert food safety authorities and the food industry where this virus could pose a non-traditional food safety risk.

## MATHERIALS AND METHODS

### XFD genomes

The XFD genomes included three human-sourced SARS-CoV-2 genomes, namely, Beijing/IVDC-01-06/2020 (EPI_ISL_469254), Beijing/IVDC-02-06/2020 (EPI_ISL_469255), and Beijing/IVDC-01-01/2020 (EPI_ISL_850948) (or briefly as IVDC-01-06, IVDC-02-06,and IVDC-01-01). The genomes were sampled at the very beginning of the XFD outbreak, one (IVDC-01-01) sampled on 2020-06-10, and the other two (IVDC-01-06 and IVDC-02-06) on 2020-06-11.

### Datasets

All the SARS-CoV-2 genomes in the background datasets were downloaded from the GISAID. We only analyzed human-sourced SARS-CoV-2 genomes with at least 29,000 bases and with complete date information.

The first dataset included the three XFD genomes and 241 genomes (referred to as domestic dataset) that were subsampled --22 genomes per division (the median genome size of divisions)-- from 893 genomes in mainland of China between 2019-12-24 (the time of the first recorted genome) and 2020-07-15. The second dataset (referred to as lineage-specified dataset) included the three XFD genomes, the five control genomes (mentioned later), and 215 genomes were subsampled (10 genomes per week as in [4]) from 14,946 genomes in the lineage B1.1.29 till 2020-07-15. The second dataset also included the earliest two genomes in Wuhan China (Wuhan/WH01/2019(EPI_ISL_406798) and Wuhan/Hu-1/2019 (EPI_ISL_402125)) as the root reference. The ending date (i.e., 2020-07-15) was chosen to keep the XFD genomes away from the boundary of the study period and to make accurate estimates of their collection dates [7].

All the genomes were processed using the Nextstrain [13] bioinformatics pipeline Augur to align genomes via MAFFT v7.4 [14] and masked spurious SNPs. The masked genomes from the Nextstrain bioninformatics pipeline (i.e., the ‘masked.fasta’) worked as input for the following leaf-dating analysis.

### Leaf-dating method

The leaf-dating method was originally developed to estimate the collection date of a particular genome with unknow collection date [7]. In this study, the expected collection dates (referred to as calculated collection dates) of the XFD genomes were estimated with leaf-dating method and were further compared with the real collection dates (or recorded collection dates) so as to provide information for inference on possible import route. This analysis was performed with the BEAST2 software [15] and used a HKY85 nucleotide substitution model with Gamma distributed rate variation and used an exponential growth coalescent model for the tree prior and assumed the strict molecular clock. The collection dates of the XFD genomes were estimated in the MCMC with a uniform prior over the period from 2020-01-01 to 2020-07-15. All the other priors used in this analysis were the same as previous studies [16]. The corresponding xml files used in the analysis were provided as supplementary files.

### Bayes factor based comparison between the expected collection date and the recorded

We introduced the bayes factor to determine if the expected collection date (denoted as Date_E_) differed from the recorded collection date (denoted as Date_r_) of a particular genome.

Specifically, we constructed two hypotheses as H_1_: Date_E_ = Date_r_ and H_2_: Date_E_ ≠Date_r_. The bayes factor of the hypothesis H_2_ over the hypothesis H_1_ (i.e., BF_21_) was calculated with the method of Savage-Dickey ratio [17] on the basis of the used prior distribution of the collection date as well as its posterior distribution generated from the leaf dating method.

### Control analysis

To further verify the validity of the analysis, we randomly selected five SARS-CoV-2 genomes from the lineage of B1.1.29 for control analysis. They were CostaRica/INC-0055/2020 (EPI_ISL_512661), England/ALDP-952FFA/2020 (EPI_ISL_553677), England/MILK-56B89D/2020 (EPI_ISL_558380), England/MILK-957274/2020 (EPI_ISL_554214), and UnitedArabEmirates/1062/2020 (EPI_ISL_698137). These control genomes had the same recorded collection date of 2020-06-11, equal to that of two XFD genomes (IVDC-01-06 and IVDC-02-06). We incorporated the control genomes into the lineage-specified dataset, calculated their expected collection dates simultaneously with those of the XFD genomes, and made comparison with their recorded collection dates based on the bayes factor.

## Data Availability

All relevant data are within the paper.

## Supporting information

**S1 Data. BEAST 2.6 XMLs and the related metadata for the two Bayesian analyses in a zipped folder**.

## Data Availability

All relevant data are within the paper.

## ACKNOWLEDGEMENTS

We appreciate helpful discussion with Dr. Tao ZHAO from Nvidia.

